# The association between sleep trajectories throughout pregnancy and postpartum pain in individuals with overweight or obesity: a prospective cohort study

**DOI:** 10.1101/2025.01.31.25320695

**Authors:** Marquis S. Hawkins, Danielle A. N. Chapa, Grace Lim, Andrea B. Goldschmidt, Michelle L. Meyer, Forgive Avorgbedor, Michele D. Levine

## Abstract

**Background:** Pain after childbirth affects maternal and infant outcomes. Although sleep influences pain in general adult populations, research on this during the perinatal period is limited. This study examines the association between sleep quality and duration changes from mid to late pregnancy and pain during postpartum hospitalization.

**Methods:** This secondary data analysis included 118 pregnant individuals (12-20 weeks gestation at enrollment) with a pre-pregnancy BMI ≥ 25 kg/m². The Pittsburgh Sleep Quality Index estimated sleep quality and duration at six prenatal visits. Group-based trajectory models identified distinct sleep patterns. Pain was assessed every 8 hours during the three-day postpartum hospitalization using a 0-10 numeric rating scale and was then calculated as the pain Area Under the Curve (AUC). Multivariable-adjusted linear regression analyzed the relationship between sleep trajectories and postpartum pain.

**Results:** Two trajectories for sleep quality and two for sleep duration were identified. The "Consistently poor" group showed increasing PSQI scores from 9 to 11, while the "Late worsening" group’s scores increased from 4 to 6. The "Late decreasing" duration group consistently slept 6-7 hours, while the "Consistently short" group maintained 5-6 hours nightly. No significant associations were found between sleep quality (expβ = 0.77, 95% CI: 0.51 to 1.17, p = 0.22) or duration (expβ = 0.75, 95% CI: 0.46 to 1.21, p = 0.24) and postpartum pain.

**Conclusions:** Sleep quality and duration changes during pregnancy were not associated with postpartum pain in this cohort. Future research should explore sleep’s impact on pain later in the postpartum period when chronic pain may develop.

## Introduction

Pain following childbirth is a postpartum recovery domain that is important to patients and clinicians.(Sultan & Carvalho, 2021) Acute and chronic pain can impede a mother’s physical recovery,(Sultan & Carvalho, 2021) affect their ability to care for their newborn,(Horvath et al., 2024; Makeen et al., 2022) contribute to poor mental health (e.g., postpartum depression),(Lim et al., 2020; Mo et al., 2022; Rajabaliev et al., 2023) and predict persistent pain.(Eisenach et al., 2008) Common non-pharmacological interventions to address chronic, pregnancy-related, or labor pain include exercise, manual therapies (e.g., massage, chiropractic, and acupuncture), and mind-body interventions (e.g., breathing exercises, muscle relaxation, and meditation).(Davenport et al., 2019; Ferraz et al., 2023; Hall et al., 2016; Hu et al., 2021; Liddle & Pennick, 2015; Santiváñez-Acosta et al., 2020; Smith et al., 2018; Zuarez-Easton et al., 2023) These interventions demonstrate moderate to large effects but have substantial heterogeneity, likely due to differences in pain experience and assessment, study design, intervention type, and population characteristics. Identifying factors explaining this variability is crucial to identifying which patients would benefit from various pain management interventions. Sleep disturbances, which can influence behavioral and cognitive aspects of pain perception, represent an essential area yet remain under-explored in the peripartum context.

The bidirectional relationship between sleep disturbances and pain severity is well-established in adults.(Rouhi et al., 2023; Varallo et al., 2022) This relationship is mediated by several pathways, including depressive symptoms, pain vigilance, pain helplessness, maladaptive coping strategies, activation of the stress system—particularly the hypothalamic-pituitary-adrenal (HPA) axis—and fatigue.(Whibley et al., 2019) There also is evidence that treating sleep disturbances can reduce pain outcomes among Veterans.(Saconi et al., 2021) However, to our knowledge, no studies have examined the relationship between sleep during pregnancy and postpartum pain.

The relationship between sleep and pain is particularly relevant in the perinatal period. Sleep disturbances are prevalent in pregnancy, often due to increasing physical discomfort, including pregnancy-related pain, alongside other factors such as hormonal changes, nocturnal awakenings for urination, and heightened anxiety related to childbirth and parenting (Christian et al., 2019; Desai et al., 2019; Hawkins et al., 2023; Munro et al., 2017; Ray-Griffith et al., 2019) Therefore, examining the relationship between sleep during pregnancy and postpartum pain can inform the development of targeted interventions, which is crucial for enhancing overall postpartum recovery. To fill the existing knowledge gaps, this study examines the relationship between changes in sleep quality and duration from early to late pregnancy and self-reported pain during postpartum hospitalization. We hypothesize that poor sleep quality and shorter sleep duration throughout pregnancy will be associated with higher overall pain during postpartum hospitalization.

## Methods

### Study Design and Population

The University’s Institutional Review Board approved all study procedures. Participants provided written informed consent or assent before enrollment. This was a secondary data analysis of a prospective observational cohort of pregnant individuals between 12-20 weeks of gestation from local obstetrics clinics from September 2012 to January 2017. The parent study aimed to assess the contributions of eating behaviors and psychosocial factors, measured repeatedly across pregnancy, to perinatal weight outcomes.(Levine et al., 2023) Participants were eligible if they 1) were ≥14 years of age, 2) had a pre-pregnancy body mass index (BMI) ≥25 kg/m2, and 3) had a singleton pregnancy. Exclusion criteria were 1) type I diabetes, 2) taking medications or being diagnosed with conditions that influence weight, 3) participating in a weight management program, or 4) reported psychiatric symptoms requiring immediate treatment. Sleep assessment (further described below) was introduced at the study mid-point (February 2015, after n=130 participants had completed the study). We included participants with complete sleep and postpartum pain data for this secondary data analysis.

### Data collection overview

The baseline assessment occurred between 12 and 20 weeks gestation (T0). It was followed by monthly assessments at 18 to 22 weeks gestation (T1), 23 to 26 weeks gestation (T2), 27 to 30 weeks gestation (T3), 31 to 34 weeks gestation (T4), and 35 weeks gestation through delivery (T5). We used data from each assessment to identify sleep trajectories.

### Sleep quality and duration assessment

The Pittsburgh Sleep Quality Index (PSQI) was used to assess sleep quality and duration. The PSQI is one of the most commonly used instruments for measuring self-reported sleep. It is highly sensitive (89.6%) and specific (86.5%) in distinguishing between good and poor sleep quality.(Buysse et al., 1989) Prior studies in pregnancy have used the PSQI to assess associations between sleep, physical activity (Hawkins et al., 2018; Tan et al., 2020), and maternal health outcomes.(Balieiro et al., 2019; Gao et al., 2019; Matenchuk & Davenport, 2020)

### Pain assessments

The primary outcome was pain intensity burden during the three-day hospitalization. This outcome was calculated as the area under the curve (AUC) based on patient self-reported pain scores ascertained clinically every 8 hours using the 0-10 numeric rating scale (0 is no pain, and 10 is the worst pain imaginable). These questions are routinely asked of patients in the postpartum unit by their nurse and recorded in the medical record. Pain intensity scores were then abstracted from postpartum medical chart records. Hospital protocol required patient pain ratings to be recorded at least every 4 hours during labor and at least every 8 hours during postpartum hospitalization (i.e., one to two days after delivery but before hospital discharge).

### Descriptive, behavioral, and clinical characteristics

Demographic characteristics, including age, relationship status (single, living with a partner, married), self-reported race, annual household income, work status (full-time, part-time, not working), education level, gestational age, and parity (number of prior births), were self-reported at baseline. Clinical characteristics, including delivery type (vaginal or Caesarean) and gestational age at delivery, were abstracted from medical records. Behavioral characteristics included smoking history (ever vs. never) and physical activity. We estimated baseline physical activity energy expenditure using the interviewer-administered Paffenbarger questionnaire. The Paffenbarger is a widely used instrument that moderately correlates with objective physical activity measures among healthy adults with overweight and obesity.(Jakicic et al., 2015) Smoking status was defined as self-reported smoking before or during pregnancy. The Center for Epidemiological Studies – Depression Scale was used to measure baseline depressive symptoms.(Radloff, 1977)

### Statistical Analysis

Descriptive statistics (i.e., means and standard deviations or N and %) were calculated for participant demographic, behavioral, and clinical characteristics. We used the Student T-test or Chi-Square to compare these characteristics between sleep quality and duration trajectory groups separately.

Using a sequential model selection approach, we employed group-based trajectory models to identify distinct trajectory groups for sleep quality and duration separately. First, we tested models with increasing numbers of groups and then evaluated the shape of the trajectories for each group (e.g., linear, quadratic, cubic). We used the Bayesian Information Criterion (BIC) to determine the number and shape of the trajectory groups, with the model yielding the lowest BIC deemed optimal. Subsequently, covariates were included to identify predictors of group membership, and those significantly associated with trajectories were included in adjusted models.

For the primary analysis, we conducted regression analyses to compare postpartum pain across trajectory groups, adjusting for covariates identified in the trajectory model or based on the team’s clinical expertise. We considered age, pre-pregnancy BMI, education, race, parity, income, relationship status, depressive symptoms, smoking status, employment status, physical activity, and delivery type (Caesarean or vaginal). Only income and depressive symptoms predicted trajectory group memberships. The teams agreed it was also appropriate to control for delivery type. Postpartum pain was transformed to address skewness in the data, ensuring the normality assumption for regression was met.

All statistical analyses were computed in the R statistical computing environment (R version 4.4.1). We used the gtsummary package (Version 1.5.2) to generate all tables (Sjoberg et al., 2021). Statistical significance was set at an alpha of less than 0.05. The statistical analysis plan was not pre-registered.

## Results

The analytic sample included 118 participants with sleep quality, duration, and pain data. For sleep duration, the two-group quadratic model had the lowest BIC value (−1269.23), indicating the best fit compared to models with additional groups or different trajectory shapes (**Supplemental Table 1**). A total of 25 people were in the first sleep duration trajectory group, which was characterized as sleeping a median of 5 to 6 hours/night throughout pregnancy. For simplicity, we called this group the “consistently short” duration group. Most participants (n=93) had a median of 7 hours/night in early/mid-pregnancy, which decreased slightly to 6 hours/night. For simplicity, we called this the “late decreasing” sleep duration group. Similarly, the 2-group linear + quadratic model was selected for sleep quality as it had the lowest BIC (−1731.32), demonstrating a superior fit compared to other models. The first sleep quality trajectory group (n=43) had a median PSQI score of 9 in early to mid-pregnancy, which gradually increased to 11 by the end of pregnancy, indicating worsening sleep quality. We called this the “consistently poor” sleep quality group. The second sleep quality trajectory group (n=75) had median PSQI scores between 4-5 in early to mid-pregnancy, which increased to 6 by the end of pregnancy. We called this the “late worsening” sleep quality group (**Figure 1**).

**Figure 1.**
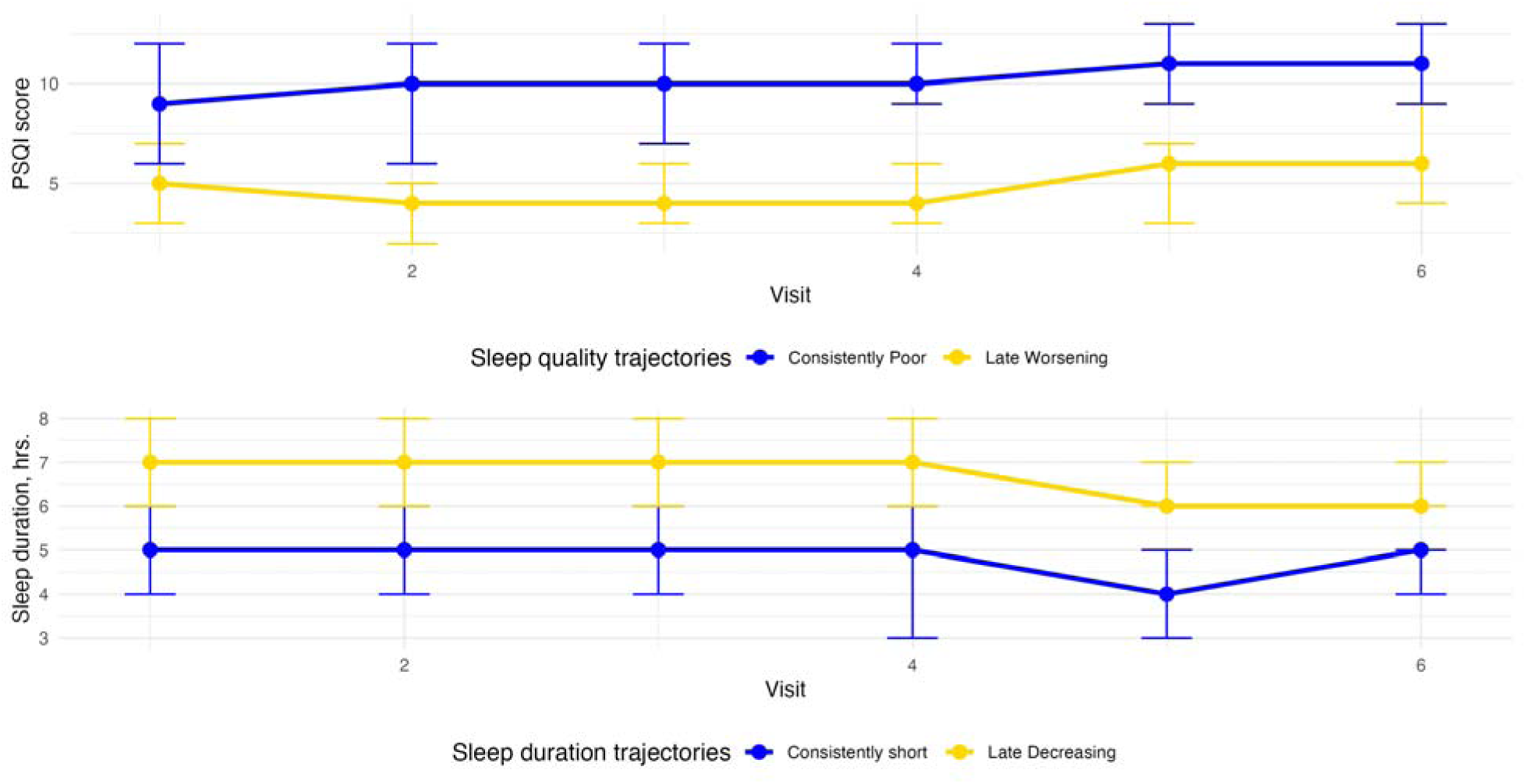
Sleep quality (top) and sleep duration (bottom) trajectory groups from early to late pregnancy. Notes: Sleep quality trajectory groups: 64% (N=75) had a “Late worsening” trajectory, and 36% (N=43) had a “Consistently poor” trajectory. Sleep duration trajectory groups: 78% (N=92) had a “Late decreasing” trajectory, and 22% (N=26) had a “Consistently short” trajectory.

Participants with consistently poor sleep quality (vs. late worsening) reported higher levels of depressive symptoms at baseline. In contrast, participants in the consistently short (vs. late decreasing) sleep duration group were younger, had higher pre-pregnancy BMI, lower levels of education, higher rates of depressive symptoms, and were more likely to be Black. Socioeconomic factors, such as income and marital status, were also more favorable in the normal to moderate sleep duration group (**Table 1**).

**Table 1.**
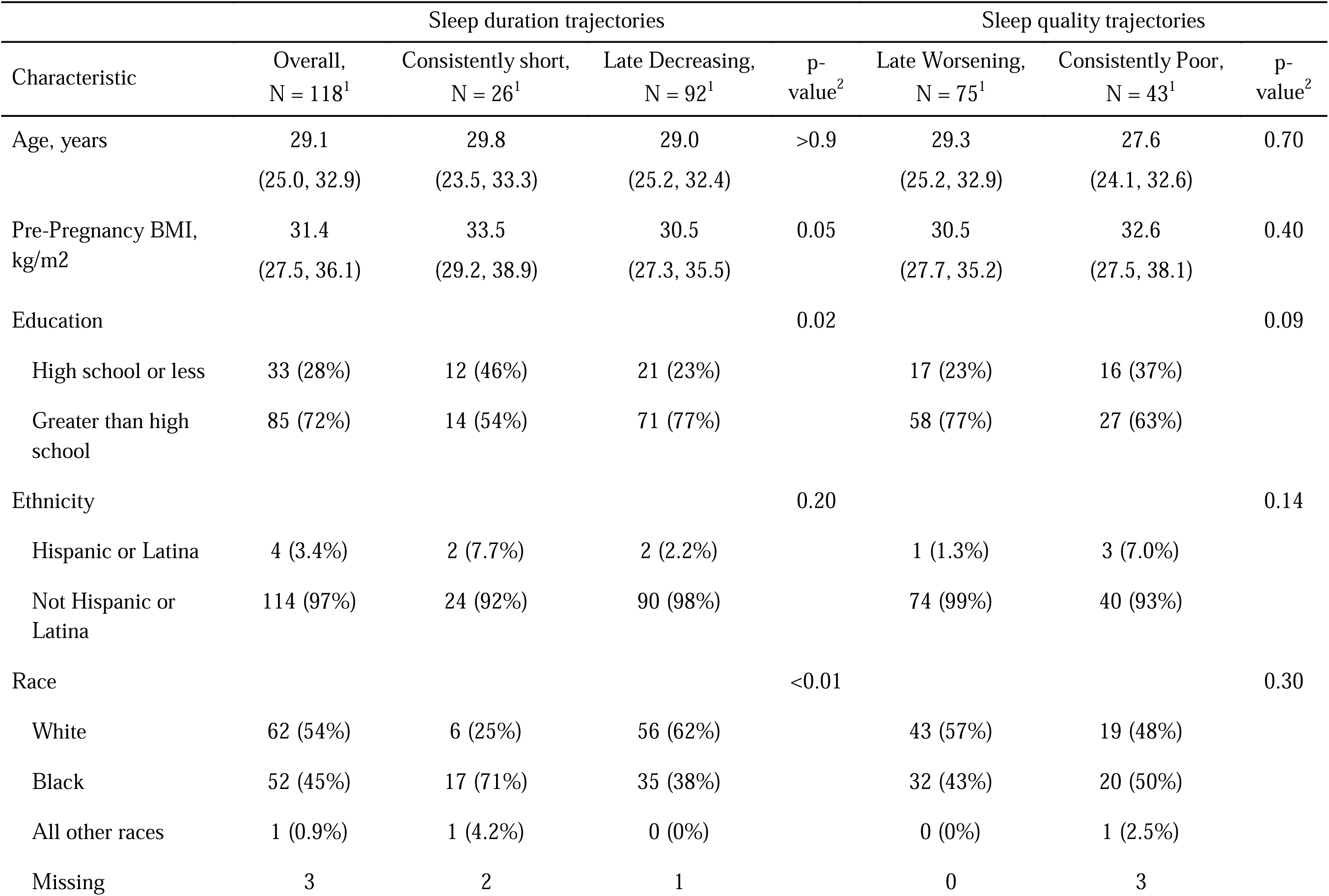
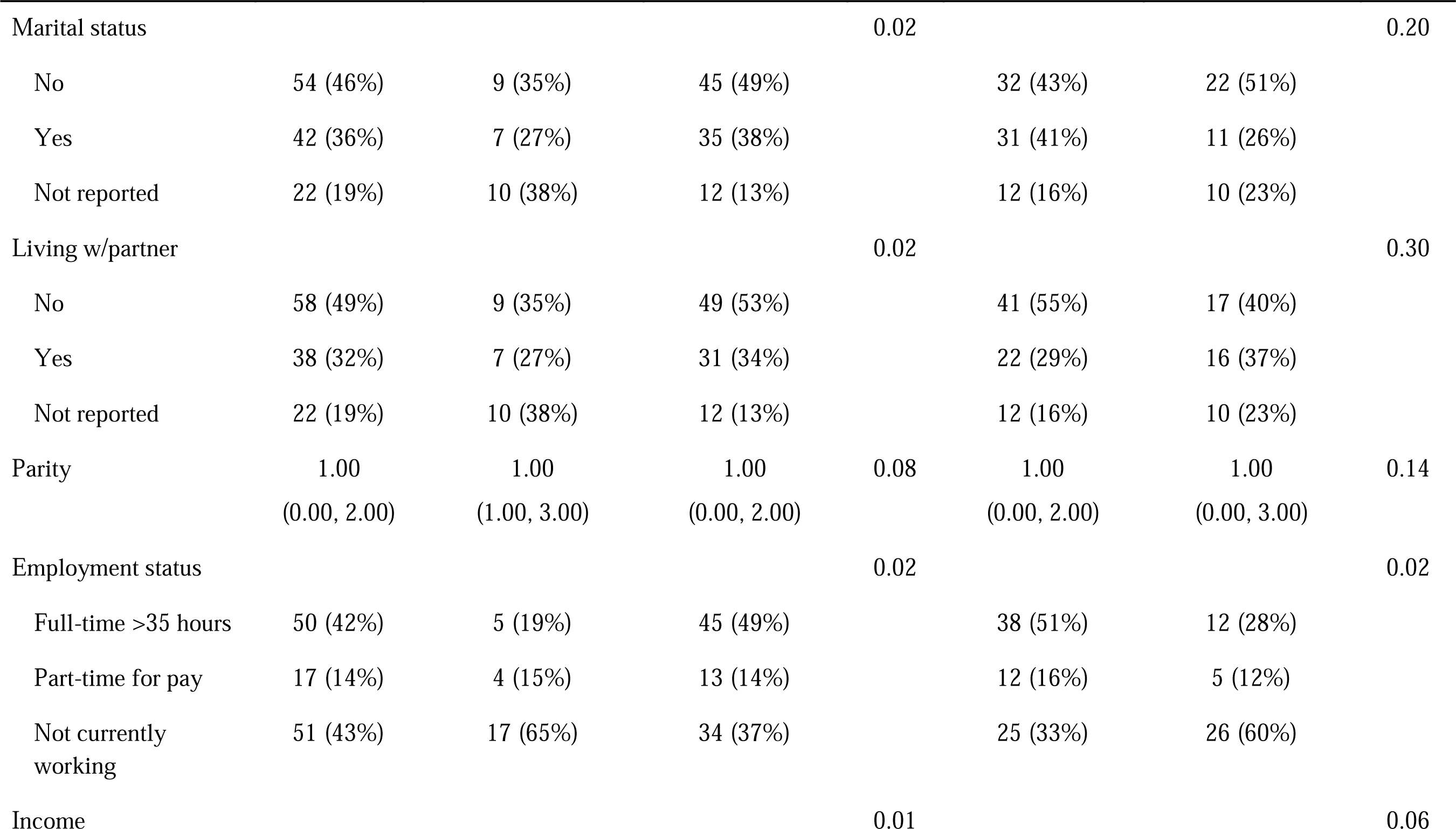
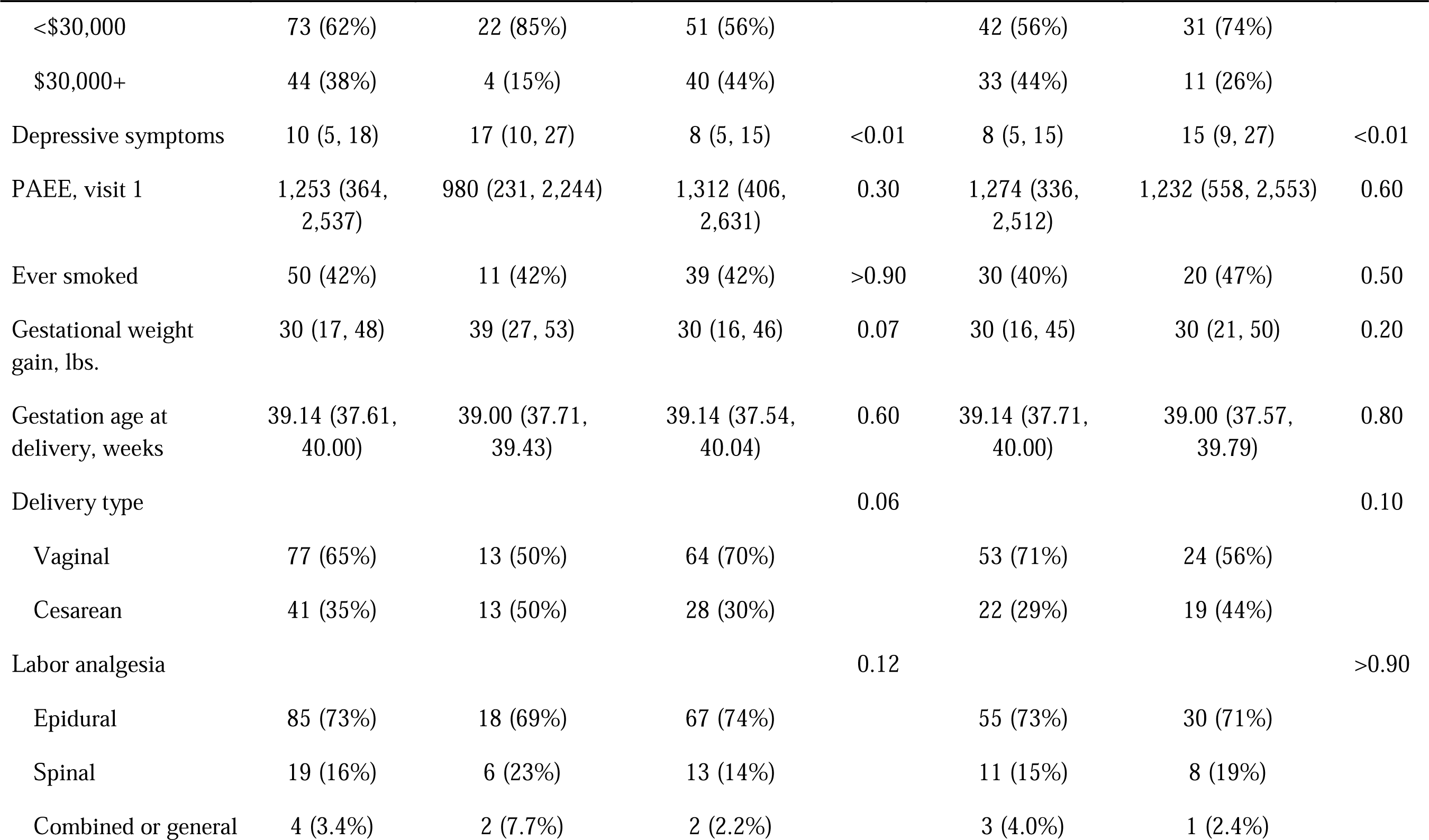
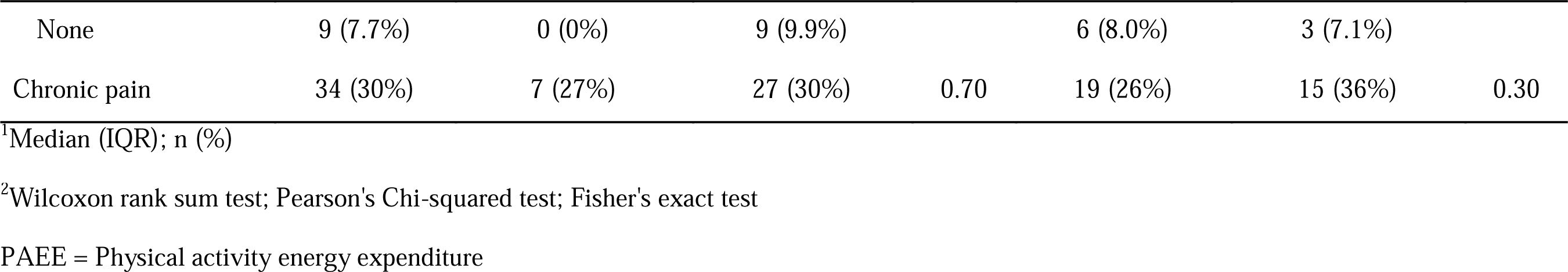
Descriptive and clinical characteristics by sleep trajectory groups.

Our primary analysis found no statistically significant differences in postpartum pain between sleep quality or duration trajectory groups (**Figure 2**). Specifically, participants who had consistently poor sleep quality had higher postpartum pain than individuals with late worsening sleep quality (expβ = 0.77, 95% CI: 0.51 to 1.17, p = 0.22). Similarly, no statistically significant difference in postpartum pain was observed between individuals with consistently short sleep duration compared to individuals with late decreasing sleep duration (expβ = 0.74, 95% CI: 0.46 to 1.19, p = 0.22). However, we are unable to rule out no or a harmful association.

**Figure 2.**
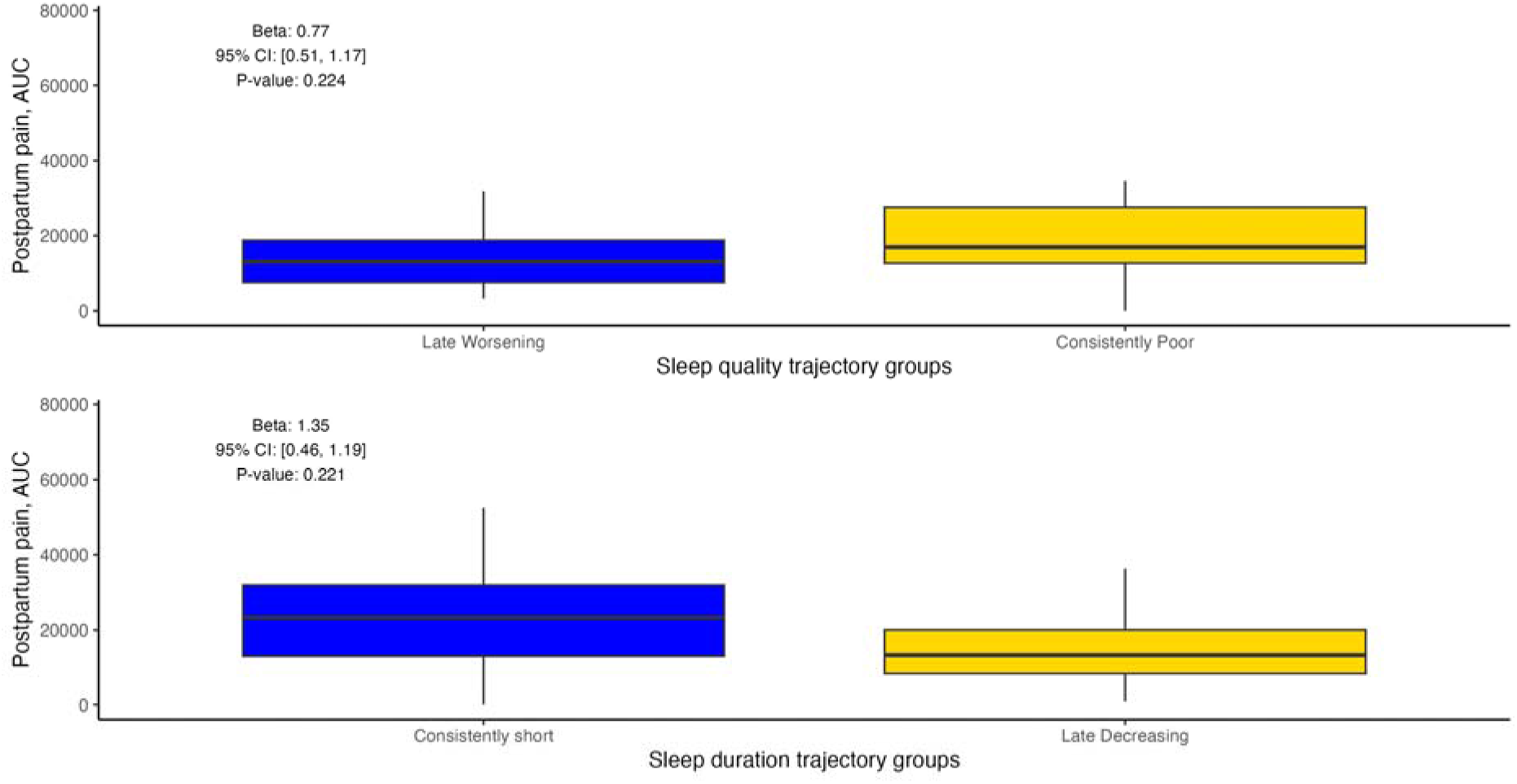
Overall postpartum pain during hospitalization by sleep quality (top) and sleep duration (bottom) trajectory groups. Footnotes: Regression models adjusted from income and depressive symptoms. Beta coefficients and confidence intervals were exponentiated to report estimates on the original scale.

## Discussion

This study examined the association between sleep quality and sleep duration trajectories from mid to late pregnancy and postpartum pain during hospitalization. We identified two sleep quality and duration trajectories, largely functions of baseline sleep quality and duration, respectively. In contrast to our hypothesis, participants with worse sleep quality and duration throughout pregnancy did not report significantly higher postpartum pain than those with relatively better sleep. These results suggest that sleep quality and duration throughout pregnancy may not directly contribute to postpartum pain levels in the immediate postpartum period among individuals with a pre-pregnancy BMI >25 kg^2^.

Our results contrast with prior studies in general adult populations, which have consistently demonstrated a bidirectional relationship between sleep disturbances and pain, with sleep disruptions often exacerbating pain perception and chronic pain conditions.(Rouhi et al., 2023; Saconi et al., 2021; Varallo et al., 2022; Whibley et al., 2019) Data among perinatal populations is limited, but the sleep-pain associations have generally been consistent with studies in general adult populations. For example, Silvertsen et al. used latent profile analysis to examine the associations of insomnia before and after childbirth and bodily pain at two years postpartum. They found that individuals with consistently high or intermediate insomnia symptoms (vs. consistently low) were associated with a nearly 2-fold increase in risk of reporting high bodily pain at two years postpartum.(Sivertsen et al., 2017) Potential explanations for our contrasting findings may be due to differences in the specific sleep domains assessed (insomnia symptoms vs. sleep duration and quality), the timing of sleep assessments (before and after childbirth vs. during pregnancy), the timing of pain assessment (two years postpartum vs. during postpartum hospitalization), or sample characteristics. In contrast to prior studies, we enrolled individuals with a pre-pregnancy BMI >25 kg/m2. Pre-pregnancy BMI is associated with poor sleep quality and shorter sleep duration during pregnancy.(Lau et al., 2022) Studies including individuals with more variability in BMI may allow them to capture greater variability and sleep to identify associations with pain.

Another explanation for the lack of significant associations could be related to the outcome measure used. While pain AUC provides a cumulative measure of pain over time, it may not capture how sleep influences pain perception at specific moments during labor or postpartum recovery. The acute pain experienced during labor and the postpartum period may be more strongly influenced by delivery-related factors (e.g., type of delivery, use of analgesics) than by chronic sleep patterns during pregnancy. Moreover, it is possible that sleep quality and duration in late pregnancy do not sufficiently impact the immediate postpartum pain experience, as most pregnant individuals commonly experience sleep disruption due to physical discomfort and hormonal changes. Future studies might benefit from assessing sleep and pain later in the postpartum period, when chronic pain may develop.

There were several limitations worth acknowledging to put the findings into proper context. The primary pain outcome was overall pain during postpartum hospitalization. Other pain measures, including self-reported quality of pain (e.g., if the pain was described as stabbing vs. debilitating) or pain tolerance, were not assessed. While labor pain was abstracted from medical records, we could not include it as an outcome due to missing data frequency. Also, we did not have pain measures later in the postpartum period, particularly chronic pain that may emerge in the weeks and months after delivery. Also, the sample size was relatively small. Therefore, we could not compare associations between sleep and pain within subgroups by race, ethnicity, or BMI groups that could moderate the association. For example, prior studies have shown that sleep disturbances during pregnancy are more strongly associated with adverse pregnancy outcomes in Black individuals than White individuals.(Blair et al., 2015) Thus, our main effects could have masked associations in subgroups. Likewise, although we observed mean differences in overall postpartum pain, they failed to reach statistical significance, which is influenced by sample size. Thus, we may have lacked adequate power to identify differences. Despite these limitations, this study has several strengths, including the prospective assessment of sleep using a validated questionnaire and pain assessment using standard procedures.

### Implications for Practice and/or Policy

This study’s findings suggest that trajectories of sleep quality and duration throughout pregnancy may not be associated with postpartum pain experienced during hospitalization. Although pain is a critical domain of postpartum recovery, this study does not provide sufficient evidence to prioritize sleep duration or quality throughout pregnancy as a target for pain management interventions during postpartum hospitalizations. These results suggest clinicians should focus on established factors and adhere to existing clinical guidelines for managing postpartum pain.

## Conclusions

In conclusion, current findings do not support a relationship between sleep throughout pregnancy and pain during postpartum hospitalization. However, the results also do not preclude a possible relationship between poor postpartum sleep and worse postpartum pain. Future research should examine long-term associations between sleep and pain after hospitalizations.

Studies should also examine mental health and socioeconomic factors, as these may play a larger role in postpartum recovery.

## Data Availability

All data produced in the present study are available upon reasonable request to the principal investigator

## Acknowledgments

The study data is not publicly available. We will provide the analytic dataset and program code with a reasonable request and data use agreement.

