## Supplemental Table 1 for "The association between sleep trajectories throughout pregnancy and postpartum pain in individuals with overweight or obesity: a prospective cohort study"

| **Supplemental Table 1** – Fit indices for identify distinct sleep duration and quality trajectory groups | | | | |
| --- | --- | --- | --- | --- |
|  | Sleep duration | | Sleep quality | |
| Model | BIC | AIC | BIC | AIC |
| **Group selection** |  |  |  |  |
| 2-group, linear | -1266.33 | -1258.01 | -1729.27 | -1720.96 |
| 3-group, linear | -1256.61 | -1244.14 | -1695.56 | -1683.09 |
| 4-group, linear | -1254.8 | -1238.17 | -1694.18 | -1677.55 |
| **Trajectory shape selection** |  |  |  |  |
| 2-group, linear | -1266.33 | -1258.01 | -1729.27 | -1720.96 |
| 2-group, linear + quadratic | -1266.98 | -1257.28 | **-1731.32** | **-1721.62** |
| 2-group, quadratic | **-1269.23** | **-1258.15** | -1725.2 | -1714.12 |

Note: Bold font indicates the final group selection
